# Impacts of weather and air pollution on Legionnaires’ disease in Switzerland: a national case-crossover study

**DOI:** 10.1101/2022.10.24.22281439

**Authors:** Fabienne B. Fischer, Apolline Saucy, Danielle Vienneau, Jan Hattendorf, Julia Fanderl, Kees de Hoogh, Daniel Mäusezahl

**Affiliations:** Swiss Tropical and Public Health Institute, Allschwil, Switzerland; University of Basel, Basel, Switzerland; Barcelona Institute for Global Health (ISGlobal), Barcelona, Spain

**Keywords:** Legionnaires’ disease, legionellosis, Legionella, weather, climate, air pollution, environmental epidemiology

## Abstract

**Background:** The number of cases of Legionnaires’ disease (LD) has risen markedly in Switzerland (6.5/100,000 inhabitants in 2021) and abroad over the last decade. *Legionella*, the causative agent of LD, are ubiquitous in the environment. Therefore, environmental changes can affect the incidence of LD, for example by increasing bacterial concentrations in the environment or facilitating transmission.

**Objectives:** The aim of this study is to understand the environmental determinants, in particular weather conditions, for the regional and seasonal distribution of LD in Switzerland.

**Methods:** We conducted a series of analyses based on the Swiss LD notification data from 2017 to 2021. First, we used a descriptive and hotspot analysis to map LD cases and identify regional clusters. Second, we applied an ecological model to identify environmental determinants on case frequency at the district level. Third, we applied a case-crossover design using distributed lag non-linear models to identify short-term associations between seven weather variables and LD occurrence. Lastly, we performed a sensitivity analysis for the case-crossover design including NO_2_ levels available for the year 2019.

**Results:** Canton Ticino in southern Switzerland was identified as a hotspot in the cluster analysis, with a standardised notification rate of 14.3 cases/100,000 inhabitants (CI: 12.6, 16.0). The strongest association with LD frequency in the ecological model was found for large-scale factors such as weather and air pollution. The case-crossover study confirmed the strong association of elevated daily mean temperature (OR 2.83; CI: 1.70, 4.70) and mean daily vapour pressure (OR: 1.52, CI: 1.15, 2.01) 6-14 days before LD occurrence.

**Discussion:** Our analyses showed an influence of weather with a specific temporal pattern before the onset of LD, which may provide insights into the effect mechanism. The relationship between air pollution and LD and the interplay with weather should be further investigated.

## 1 Introduction

Legionnaires’ disease (LD), caused by inhalation or aspiration of the bacteria *Legionella* spp., is a severe form of pneumonia with a high case-fatality rate of 10%^1^. *Legionella* spp. are ubiquitous in the environment, particularly water, and grow optimally in stagnant, warm water (25 to 42 °C)^2^. Therefore, most water reservoirs that aerosolise or evaporate are potential risk sources for infection^3^. Numerous infectious sources have been reported ranging from residential drinking water ^4^, cooling towers, whirlpools, potting soil/compost, to fountains and wastewater treatment plants. Evidence stems mostly from outbreak investigations. However, the largest part of all LD cases are community-acquired and not related to an outbreak. As such, the main sources of infection for sporadic cases remain unknown^5^.

LD case numbers have been increasing in many countries, where the disease is surveilled. In the EU the notification rate increased from 1.4 cases per 100,000 population in 2015 to 2.2 in 2019^6^. The US reported an increase from 1.9 to 2.7 per 100,000 population between 2015 and 2018. The reason for this widespread increase is unclear. Apart from improved disease surveillance, the design and maintenance of building infrastructure, and an ageing and increasingly susceptible population, Barskey et al. suggest that the geographical distribution and increasing seasonal frequency of cases in summer indicate weather patterns may play a role in the increasing LD incidence^7^. Studies from the European Centre for Disease Prevention and Control (ECDC) and others also consider climate change as one of the potential drivers of the increasing temporal trend^6,8^.

Weather has been strongly linked to the incidence of LD^8,9^. Precipitation, high relative humidity^10-16^ and warm temperatures^12,14,16-20^ before the disease onset were often reported as important risk factors. Other relevant risk factors that were less often identified included atmospheric pressure^13,19^, low wind speed^10,13^, high dew point and low daily visibility^13^. High vapour pressure was reported as a significant risk factor by Conza et al.^20^. Yet, temperature, relative humidity and vapour pressure are all closely interlinked and most studies included relative humidity in their investigation rather than vapour pressure. Most of the studies faced similar limitations: Vapour pressure and temperature are almost perfectly correlated strongly hampering disentangling individual association with LD incidence. In addition, most of them had to accept limitations in spatial resolution, as the exposure data came from a limited number of weather stations and/or were averaged over a larger area. Lastly, the variable incubation period of LD of 2 to 14 days^21^, together with the high correlation between consecutive days’ temperature and similar weather variables, make it difficult to select an appropriate timeframe for which an association with weather may be relevant. Most studies either calculated the odds ratio separately for each day or calculated the odds ratio averaged over a given time window. While the a priori definition of these windows can affect the observed associations, these models are also subject to exposure misclassification and autocorrelation^10^.

Despite the evidence that ambient air pollution has both short- and long-term effects on respiratory health^22,23^ and the risk of respiratory infections^24^, the role of air pollution in LD incidence has received little study, both, alone and in association with weather^22,23^. One previous study investigated the short-term impact of particulate matter (PM) on LD in Portugal and attributed part of a larger LD outbreak to a Saharan dust storm that yielded high PM_10_ concentrations and favoured aerosol formation^25^. As ambient air pollution is a complex mixture of compounds including PM and volatile pollutants, such as NO_x_ or ozone (O_3_), it is often difficult to disentangle the harmful effects of different pollutants individually. Similarly, the strong association between weather and air pollution makes it difficult to estimate interaction and causal associations concerning LD infections^26,27^.

In Switzerland, LD is included in the national surveillance system for infectious diseases and case numbers doubled in the last decade reaching approximately 560 cases in 2021^28^. Similar to other countries, the cause for the increase remains widely unknown, but distinct epidemiological features such as a pronounced seasonality with most cases occurring between June and September are observed^29^. There is also a clear regional distribution of cases in Switzerland^29^ with the southern canton of Ticino constantly reporting the highest notification rates in the country. The reason for this divergence is unclear, yet studies on the positivity rate and physicians’ testing behaviour suggest that this is not due to a difference in testing and reporting behaviour alone^30,31^. In Switzerland, the Alps act as a barrier between the South and the North of the country. Hence, the regions north of the Alps are influenced by the Atlantic Ocean resulting in mild, humid winters and drier summers, while the southern region is influenced by the Mediterranean Sea resulting in even milder winters and warm and humid summers. One study investigated the difference in incidence and weather between Ticino and a region north of the Swiss Alps and found that higher vapour pressure in Ticino significantly increased the risk of developing LD^20^. While air pollution has strongly decreased in Switzerland since 1985^32^, ground level concentration limits are still surpassed regularly, particularly for tropospheric ozone^32^. The daily thresholds for PM are also exceeded multiple times each year, and the highest measurements are being recorded in Ticino. Gaining evidence on the potential role of air pollution in LD incidence is therefore particularly relevant to inform future public health policies.

The aim of this study was to understand the role of environmental factors on LD incidence in Switzerland. We conducted a series of comprehensive analyses exploiting the Swiss LD notification database in conjunction with detailed spatial data to: 1) understand the spatial distribution of LD cases at cantonal and district levels and identify spatial clusters of LD; 2) elucidate the ecological determinants of LD and 3) study the association of short-term weather and air pollution on LD incidence. The latter used a case-crossover design to achieve high temporal and spatial resolution across the whole of Switzerland. An overview of the analytical approaches is depicted in Figure 1.

**Figure 1.**
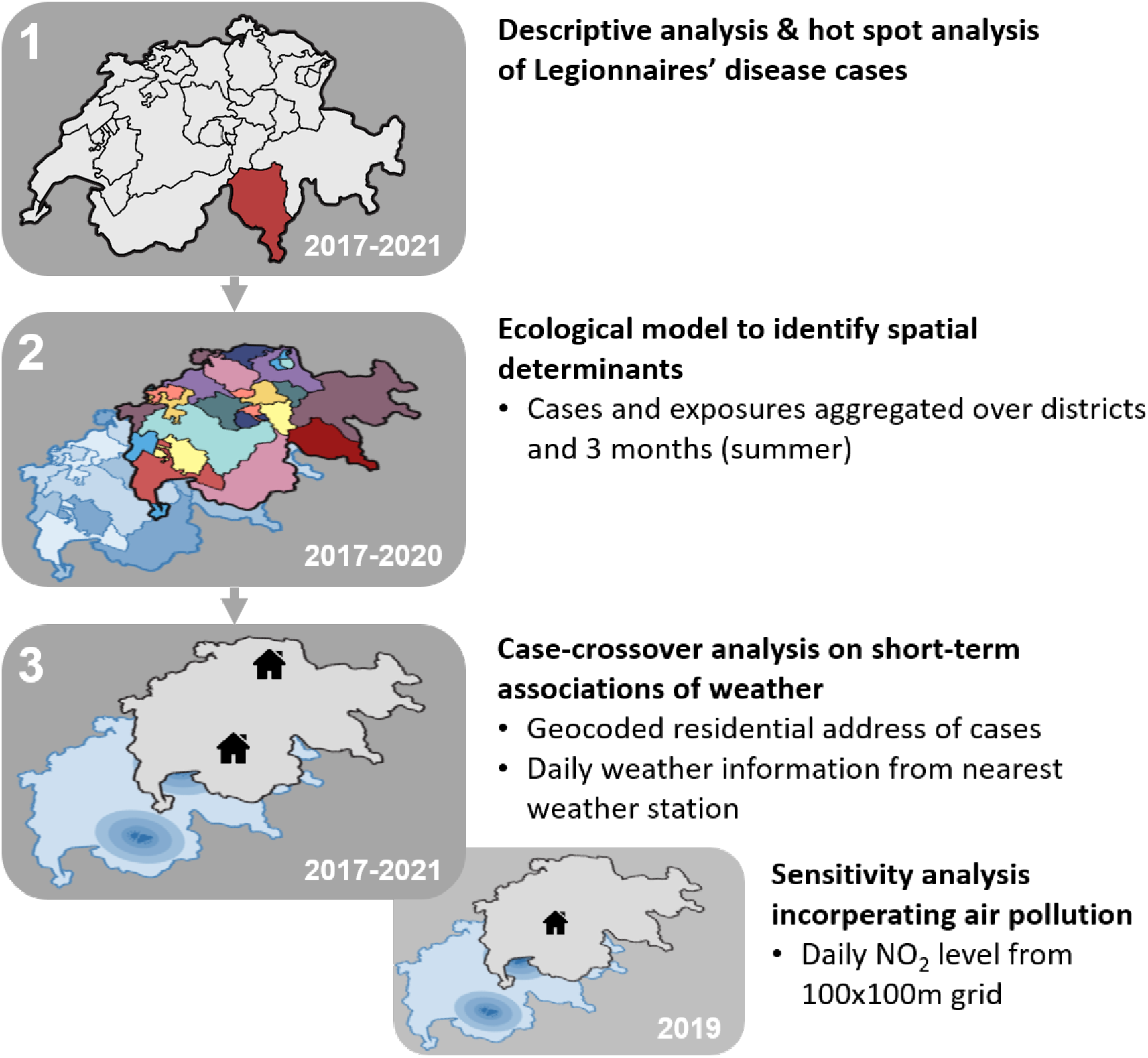
Overview of the analytical approaches presented in this paper. All analysis were based on the Swiss national notification data for Legionnaires’ disease from 2017 to 2021. The first analyses comprised of a descriptive and hot spot analysis. The second analyses was an ecological regression model using various environmental exposures (such as wastewater treatment plant locations or degree of urbanisation). The third analysis was a case-crossover analysis on the short-term association of weather with LD cases. The third study incorporated a sensitivity analysis restricted to 2019 but incorporating air pollution data (daily NO_2_ levels).

## 2 Methods

### 2.1 Study design, Legionnaires’ disease notification data sources, access and processing

This is a longitudinal retrospective study utilising routinely health data for LD collected from the National Notification System for Infectious Diseases (NNSID) in Switzerland. We considered disease notifications from 1 January 2017 to 19 November 2021. We applied the case definition of the Federal Office of Public Health (FOPH)^33^ and included only confirmed and probable cases for the analysis limited to community-acquired LD cases; hence, travel-associated, nosocomial and occupation-associated LD cases were excluded. Further, we excluded all cases with residency outside of Switzerland and cases with missing demographic information (age and sex). Residential information was geocoded using the geocoding tool of the Federal Office of Topography (swisstopo)^34^, and cases without known residency at district-level were excluded.

### 2.1 Exposure data

#### 2.2.1 General spatial/environmental determinants

For the ecological model, we compiled the geolocations of the following suspected LD exposure sources and aggregated the exposure levels by district: freshwater bodies and streets, air pollution and weather, wastewater treatment plants, composting plants, socio-economic position and age of the population, degree of urbanisation, land use and population density (Table S1).

#### 2.2.2 Meteorological and air pollution data

Meteorological data were obtained from the Federal Office for Meteorology (MeteoSwiss) for daily mean air temperature 2 m above ground, daily mean relative air humidity 2 m above ground, daily total precipitation, daily mean vapour pressure 2 m above ground, daily mean wind speed (scalar), daily maximal gust peak (one second), and daily mean atmospheric pressure at barometric altitude (QFE). Data were obtained for 191 weather stations in Switzerland for the timeframe from 1 November 2016 until 19 November 2021. Meteorological data were checked for implausible values and outliers. Outliers were defined as measured values that deviated more than a predefined cut-off value (e.g. 20 °C for temperature) from predicted values. The prediction models were based on spatial coordinates, altitude and the daily median of the respective weather parameter across all weather stations included in the study as fixed effects and the site id of the weather station as random intercept.

We omitted monitoring stations with less than 75% data availability for precipitation and 80% data availability for all other weather variables. For the remaining stations, missing daily values of each variable were imputed using information from the meteorological stations with complete data (Table S2). To impute missing daily exposure values, we fitted separate linear regression models for each monitoring station and variable using daily values from all stations with complete data and including month, year and Julian day as fixed effects (Equation 1). The models’ performance was assessed by calculating the R^2^ and adjusted R^2^ comparing the imputed and measured variable values. In addition, temporal evolution of the imputed values were visually assessed for each monitoring station (example in Figure S1). Finally, all missing weather values were replaced by the imputed values.

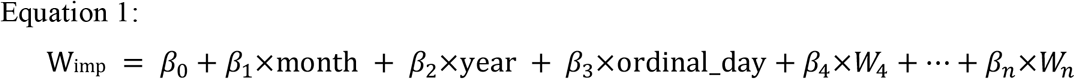

Where W_imp._ is the imputed weather value, and W are the weather values at monitoring stations with complete data.

All weather estimates were extracted at the home location of LD cases, defined as the values from the weather station closest to their home location for each weather variable individually. If the altitude difference between the home location and weather station was over 500 m, the second-closest weather station was selected. Only, if the second-closest station also had more than 500 m in altitude difference, the closest station remained selected.

Mean daily NO_2_ concentrations were extracted for each case’s home location from the spatio-temporal model estimating historical NO_2_ concentrations in Switzerland at a fine resolution (100 × 100 m; daily estimates from 2005-2016, and 2019) described by de Hoogh et al.^35^. As the address-level information for LD cases was only available for 2017-2021, we could investigate the association between NO_2_ concentration and LD incidence only for the year 2019.

### 2.3 Statistical methods

#### 2.3.1 Descriptive and hot spot analyses

Data content and data quality were analysed descriptively using the statistical software R Version 4.0.3^36^, Stata Version 16^37^ and ArcGIS Version 10.6.1^38^. Notification rates, defined as the number of notified cases per 100′000 resident population, were calculated using publically available population statistics from the Federal Statistical Office (FSO)^39^. We used the Pearson correlation to assess correlation between the weather variables. P-values <0.05 were considered statistically significant.

Two global statistics (Getis-Ord General G and Global Moran’s I) and two local statistics (Getis-Ord Gi* and Local Moran’s I) were used to identify hot spots of sex- and age-standardised notification rates for LD on district level (n= 143). Relationships between districts were determined via ‘zone of indifference’, and distance bands selected using ‘Incremental Spatial Autocorrelation’ to ensure each district had at least one neighbour. The Local Moran’s I analysis was conducted with 9’999 permutations. A false discovery rate (FDR) correction accounted for multiple testing in both local analyses.

#### 2.3.2 Ecological model

An exploratory analysis using an ecological model was performed to investigate the association of LD case frequency with general spatial/ environmental determinants. We used univariable and multivariable negative binomial regression models to explore associations between LD case counts per district (adjusted for the population size) and exposure source densities (for count data; e.g. infrastructural exposure sources) or values (e.g. mean age of the population) using the log-transformed population size as offset (Table S1). Separate models were developed for PM_2.5_ and NO_2_. All models were adjusted for the number of compost facilities and wastewater treatment plants, length of shoreline and rivers per district. As well as the mean age, mean PM_2.5_ and NO_2_ levels, mean temperature, mean relative humidity and mean precipitation per district and land coverage ratio and category of degree of urbanisation and Swiss socioeconomic position (Swiss-SEP).

#### 2.3.3 Case-crossover design

The ecological model described above is based on aggregated data at the temporal and district levels, which help inform about relevant characteristics of the regions with increased LD incidence rates and potential environmental candidates that can drive these differences. However, remaining bias resulting from regional differences not captured in the models cannot be avoided with this approach. To address this issue, we used LD infection data at the individual level and conducted a case-crossover study investigating the short-term impact of a set of meteorological (and air pollution) exposures at fine spatial and temporal resolution. The case-crossover is a self-matched study design where each exposure levels during the “hazard period” (the period before the adverse outcome occurred) is compared with exposures in other periods where the case did not occur (disease-free period) (Figure 2). The self-matching procedure limits the risk of potential confounding by time-invariant characteristics (e.g. sex, socio-economic position, chronic comorbidities and other unknown regional confounding suggested by the high heterogeneity in regional notification rates), which are typical sources of bias in other observational study designs and ecological studies.

**Figure 2.**
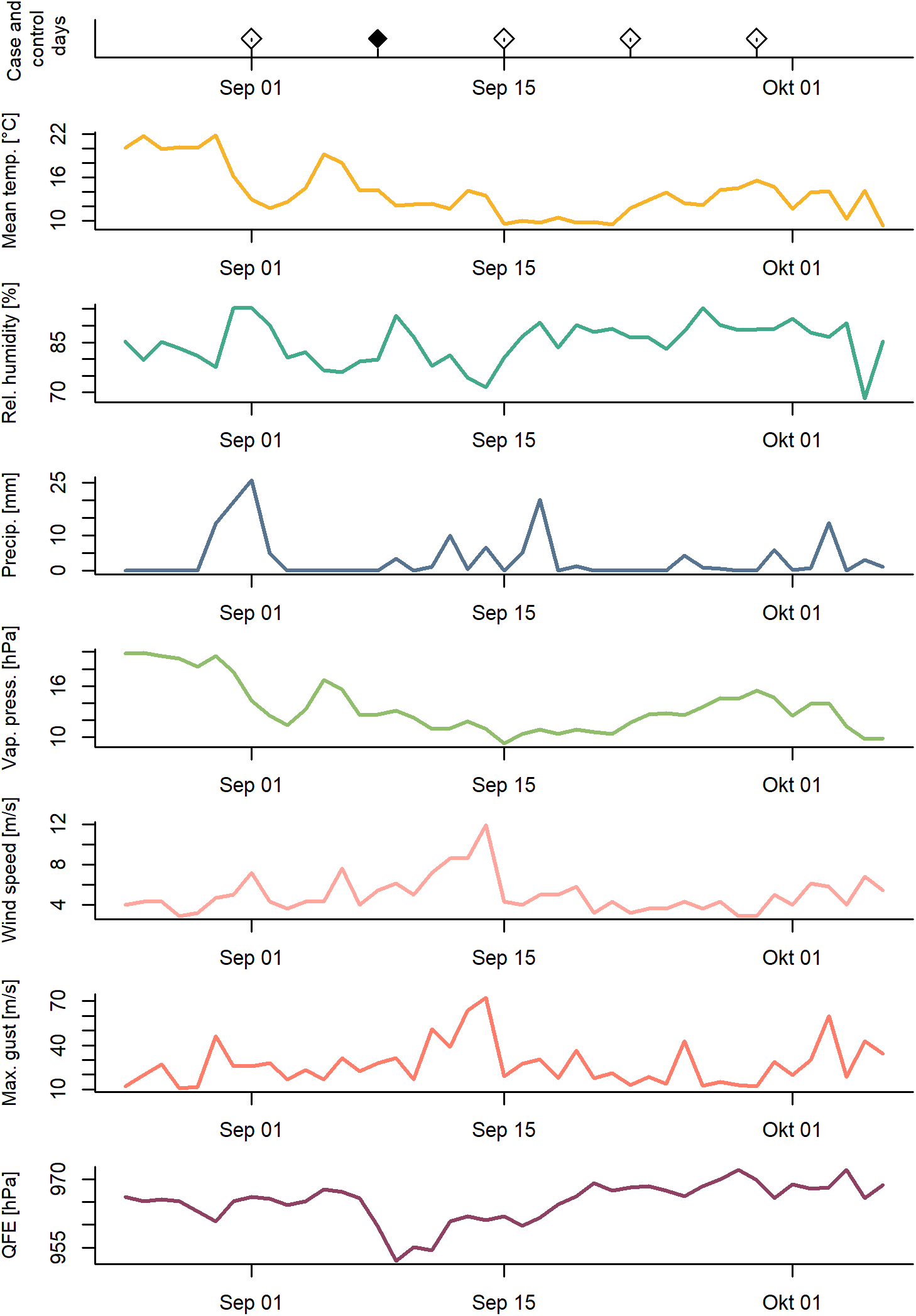
Illustrative example of the time-stratified case-crossover study design and the data. The first panel shows the case (black) and three control windows (white), which were chosen randomly before or after the case within the same month, for one individual. The lower panels show the time series of the weather at the residential address of this case.

To avoid any risk of bias due to seasonality and time-trends, the control periods were selected using a time-stratified sampling approach matched on the same day within the same month, leading to four to five control events for each case event^40^.

We conducted single- and multi-exposure conditional logistic regression to estimate the association between individual weather exposures and the risk of infection by *Legionella*. First, we fit single-exposure distributed non-linear lag models (DLNMs) to estimate the association between single exposures and the risk of infection up to 21 days prior to the onset of clinical manifestations (using the R package dlnm^41,42^). These lag periods were chosen as 21 days are a typical lag used when analysing temperature and weather effects on hospital admission and mortality^43,44^. Further, it captures the typical incubation days for LD of 2 to 14 days^21^.

The lag function was specified as a natural spline with one to three equally spaced knots on the logarithmic scale. The exposure was specified as a linear term (precipitation, gust, atmospheric pressure, relative humidity); mean temperature as a b-spline with two knots (50^th^ and 75^th^ percentiles of the annual distribution) and vapour pressure as a b-spline (1 knot at the median of the annual distribution). The best combination of lag and exposure function was determined by Akaike’s criterion. Given the high correlation between mean daily temperature and mean daily vapour pressure (Pearson’s correlation r= 0.90), we constructed two separate multi-exposure models. All models were adjusted for regional school holidays, defined as the total number of days of holidays during the incubation period to account for travelling. We did not add a variable adjusting for the effects of the COVID-19 pandemic, as seasonal time-trends are naturally accounted for by the study design.

For easier interpretation, we estimated the odds ratio (OR) of infection as the ratio between the odds at 0 and the odds at a sensible value: for mean temperature 20 °C, precipitation 10 mm, wind speed 20 m/s and maximal gust 20 m/s. For variables that only take positive values (atmospheric pressure, mean relative humidity and vapour pressure), we estimated the OR as the deviation from the median to the 5^th^ and 95^th^ percentiles of the annual distribution. To disentangle the role of environmental and weather conditions on different phases of the disease transmission, we present the overall odds ratios for three a priori selected exposure windows: ‘Early incubation/ shortly before disease onset’ (lag 2-6), ‘Prolonged incubation period’ (lag 6-14) and ‘Before incubation’ (lag 14-21).

#### 2.3.4 Sensitivity analyses

We conducted the following three sensitivity analyses: (1) to validate the estimates from our DLNM models, we built ‘simple’ models using each weather variable’s average over the most relevant lag days (as identified by the DLNM) as exposure variable and conducted single and multi-exposure conditional logistic regression adjusted for school holidays. Further, these simple models provided estimates of the variation inflation factor and overall correlation between the different exposures, and helped inform and verify the validity of our DLNM multi-exposure models. (2) Air pollution can change rapidly over time and is partly correlated with meteorological conditions, making it a possible confounding factor for our analyses. To rule out this potential bias and explore the effect of air pollution on LD, we considered data from 2019 for which individual daily NO_2_ estimates were available. We compared our results from the DLNM and ‘simple’ models with and without additional adjustment for NO_2_; and, (3) Ticino has reportedly the highest LD notification rates and unique weather conditions. To ensure that no other time-variant factors special to Ticino bias our results, we ran a sensitivity analysis excluding all cases from Ticino.

### 2.4 Ethical approval

The study was conducted under the Epidemics Act (SR 818.101)^45^. The study was submitted to the Ethics Committee Northwest and Central Switzerland (EKNZ), and was evaluated to be outside the scope of the Human Research Act (SR 810.30)^46^ and, therefore, does not require ethical approval.

## 3 Results

### 3.1 Legionnaires’ disease cases 2017-2021, description and hot-spot-analysis

Between 2017 and 2021, 2,854 cases of LD were reported in Switzerland. We excluded 376 cases with a ‘possible’ (N=151) or missing case definition (N=135), and 405 cases due to being categorised as ‘travel-associated’, ‘nosocomial’ or ‘occupation-associated’. An additional 20 cases were excluded for the following reasons: missing sex (N=1), district (N=11) or non-Swiss residency (N=8). Six cases occurred after 19 November 2021 and were, thus, excluded from the analyses. In total, 2,047 cases of LD with an onset between 1 January 2017 and 19 November 2021 were included in the study (Table 1). Among the cases, 68.5 % were males and 84.9 % were over the age of 50 years. Most cases occurred in the summer months between June to August (40.3 %).

**Table 1.**
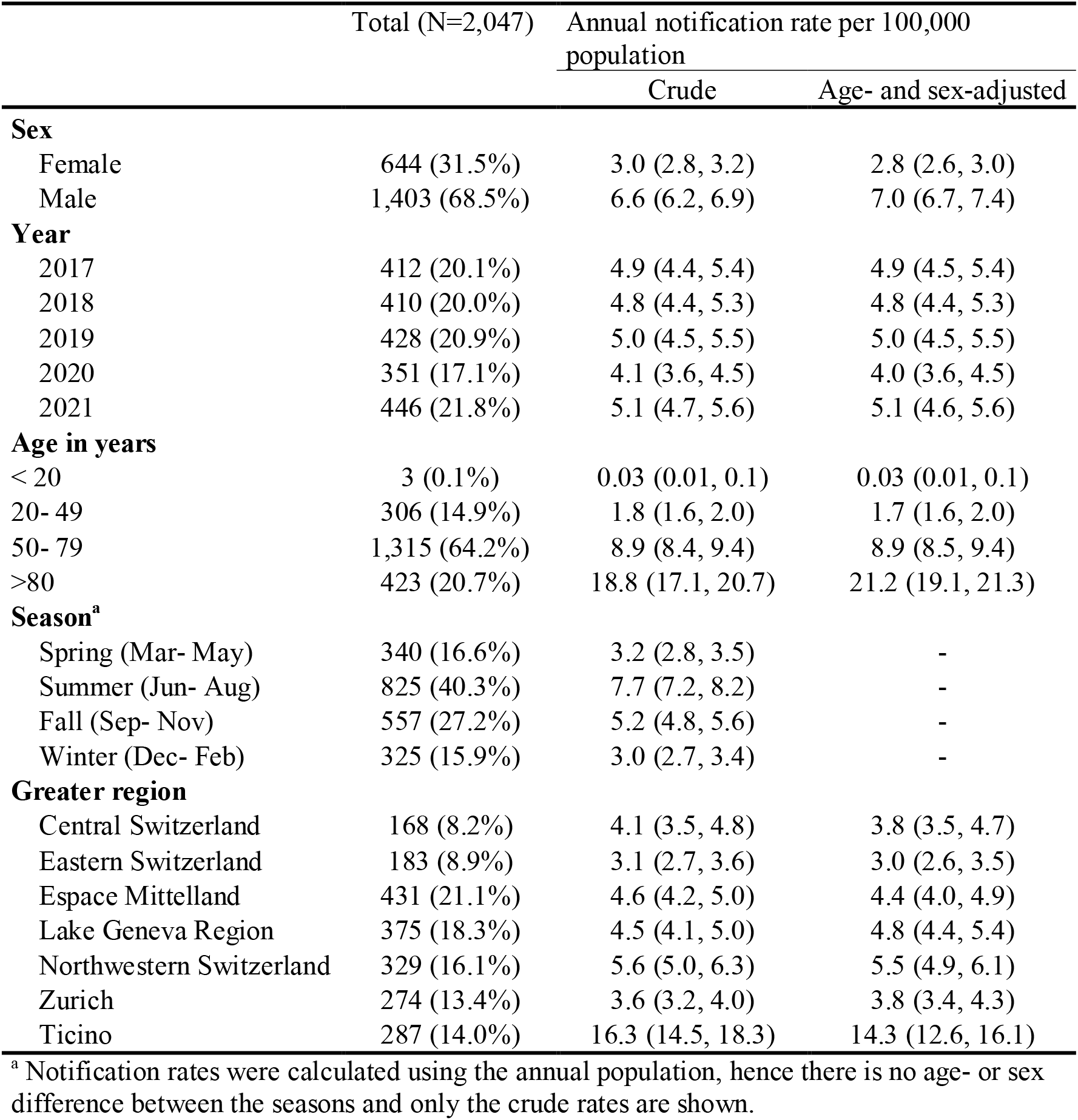
Legionnaires’ disease cases and annual crude and age-and sex adjusted notification rates in Switzerland, 2017 - 2021

The annual age-and sex adjusted notification rates were highest in the greater region (NUTS-2 level, N=7 in Switzerland) of Ticino with 14.3 cases/100,000 population (CI: 12.6, 16.0). The lowest notification rates were observed in the region of Zurich (3.8, CI: 3.3, 4.3). In the canton of Ticino, the district of Lugano (20.6, CI: 17.7, 24.0) had a 33% higher notification rate than the district of Mendrisio with the second highest notification rate (14.0, CI: 10.0, 19.3) (Figure 3A and B). Hot spot analyses, with the two local statistics Local Moran’s I and Getis-Ord Gi*, both showed a significantly elevated notification rate in Ticino (Figure 3C and D).

**Figure 3.**
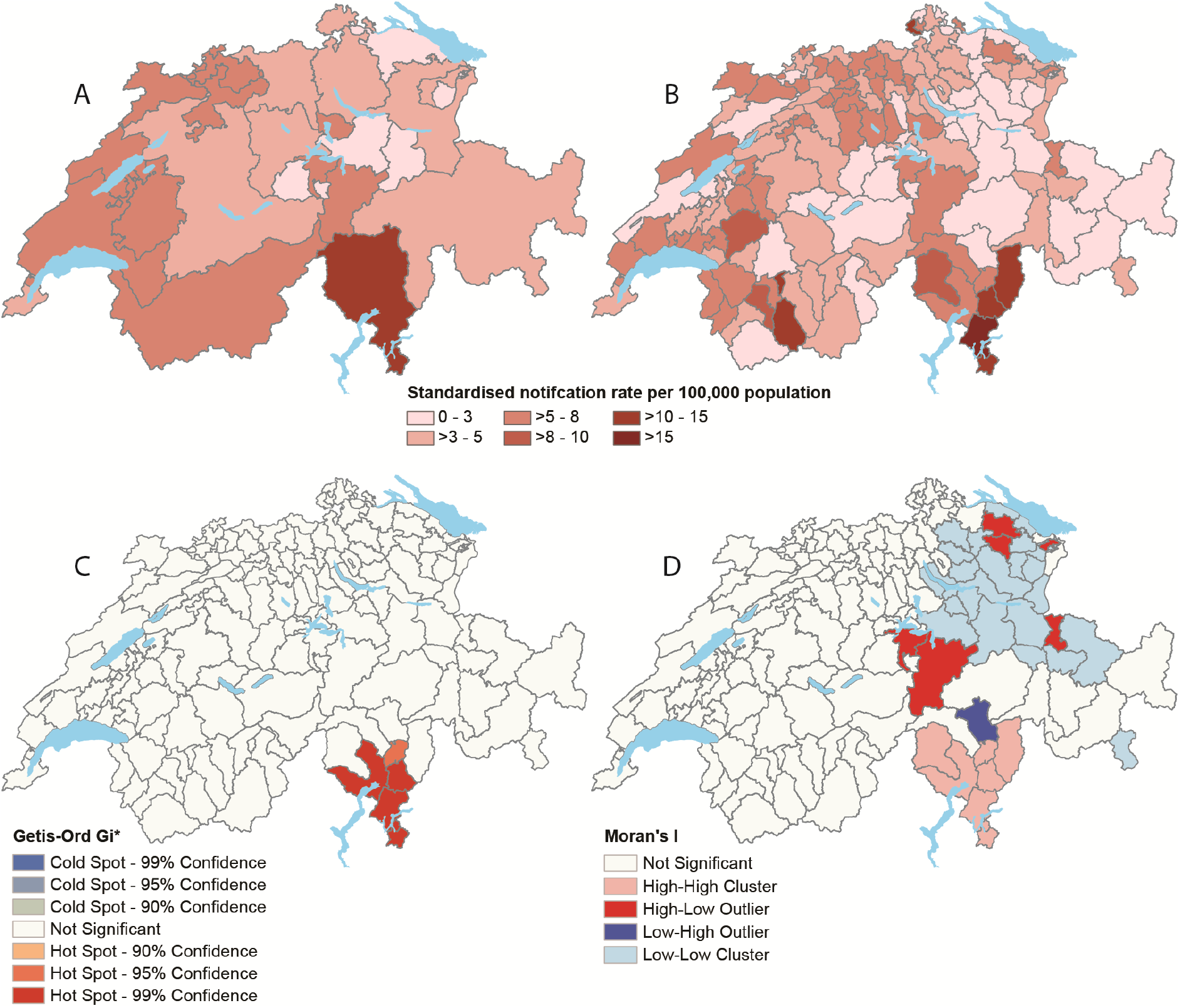
Distribution of Legionnaires’ disease cases in Switzerland from 2017 to 2021. **(A)** Average annual notification rate across cantons. (**B**) Average annual notification rate across districts. (**C**) Cluster analysis using Local Moran’s I. (**D**) Cluster analysis using Getis-Ord Gi*.

### 3.2 Ecological model to identify spatial determinants (2017-2020)

We included 1,603 LD cases between 2017 and 2020. Since both PM_2.5_ and NO_2_ were statistically significant in the univariable model, and there is limited existing literature on the association of air pollution and LD occurrence, we developed one separate model for each air pollutant. Results from both multivariable regression analyses suggest that the relative humidity is negatively associated with LD occurrence (Figure S3). Higher air pollution concentrations (PM_2.5_ and NO_2_) were strongly associated with LD occurrence. Further, lower socio-economic position and older age were positively associated with LD occurrence on a district-level. Infrastructural and environmental exposures (e.g. wastewater treatment plants, lakes) were not found to be associated with LD case numbers, but effects could have been masked by aggregation across a larger area.

### 3.3 Case-crossover analysis on short-term determinants

#### 3.3.1 Weather in Switzerland (2017-2021)

Weather data were linked to cases’ individual locations. For most cases (88.5%), a residential address was reported and used. If not available (11.5%), the geometric centroid of the reported residential municipality was used as address.

From the data of the seven weather variables of the 191 stations, only one value was implausible and set to missing. On average, the imputation models for missing weather data performed well (0.91 adjusted R^2^) and only a few stations for the variables “gust” and “maximal relative humidity” scored an R^2^ lower than 0.6 (**Error! Reference source not found**.). The median distance and median altitude difference of individual cases to the linked station was 5.4 km (range 0.1 to 32.6 km), respective 37 m (range: 0 to 630 m) (Figure 4).

**Figure 4.**
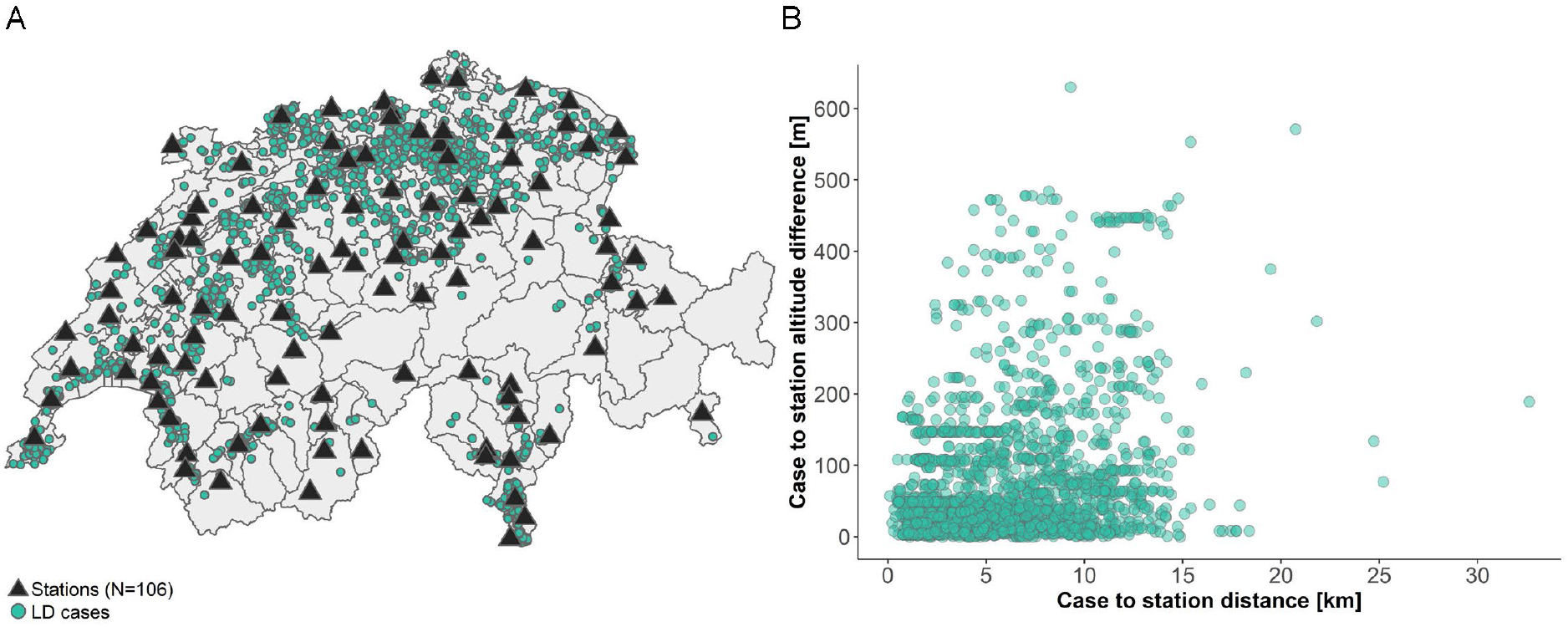
Assignment of weather exposure to Legionnaires’ disease cases (2017-2021) on the example of mean daily temperature. (**A)** Map of all included Legionnaires’ disease cases (2017-2021) in green and included weather stations measuring temperature (black triangles). (**B)** Linear distance and altitude difference of each case to the selected weather station measuring temperature.

In general, temperatures are cooler in the alpine regions all-year. Across Switzerland, Ticino is subject to most heavy rain events. The year 2018 was exceptionally warm with heat periods above 30° C across Switzerland and notable lack of rain during summer. The year 2019 had an equally hot summer but with more precipitation. The year 2020 was characterised by a mild winter, another hot summer and heavy rain events in Ticino (August and October) and the Lake Geneva region, Berne, and parts of Graubünden (October only). In 2021, June and July were exceptionally wet across Switzerland with heavy rainfall in Ticino and several flooding north of the Alps. **Error! Reference source not found**. in the Supplementary Material provides an overview of the weather conditions from the stations included in our study.

At address-level resolution, daily values of (1) mean vapour pressure and mean temperature and (2) gust and wind speed (r = 0.79) were strongly correlated with each other (Pearson’s correlation r = 0.90). Weaker correlated were mean temperature and mean relative humidity (r = -0.31), atmospheric pressure and vapour pressure (r =0.31), and precipitation and relative humidity (r = 0.31).

#### 3.3.2 The association between weather and Legionnaires’ disease case frequency (2017-2021)

We estimated the risk of *Legionella* infection for seven weather variables using single-exposure DLNM models (Table 2). Six of these variables were included in a multi-exposure model which also included either mean temperature (Model 1) or mean vapour pressure (Model 2), due to their high collinearity. We validated the models running simple conditional logistic regressions over either the statistically significant lag days or meaningful lag periods if no significant lag days were observed (wind speed and maximal gust: 1-5 days) (Table S4 and Figure S4). Overall, the results of the DLNM and simple models were consistent.

**Table 2.** Output for DLNM models using conditional logistic regression. Odds ratios and 95% confidence intervals for single-exposure and multi-exposure models for each weather variable. Due to collinearity, two models were constructed, once with temperature and the other with vapour pressure. All estimates stem from the mean temperature model (Model 1), except vapour pressure, which is based on the vapour pressure model (Model 2). The centre depicts the reference value selected for the prediction. The value depicts the value for which the overall odds ratio are estimates.

| Variable | Centre | Value | Lag period | Single-exposure |  | Multi-exposure |  |
| --- | --- | --- | --- | --- | --- | --- | --- |
|  |  |  |  | OR | 95% CI | OR | 95% CI |
| Temperature | 0 °C | 20 °C | 2-6 days | 0.91 | (0.59, 1.42) | 1.10 | (0.69, 1.74) |
|  |  |  | 6-14 days | **1.93** | **(1.20, 3.10)** | **2.83** | **(1.70, 4.70)** |
|  |  |  | 14-21 days | **1.77** | **(1.13, 2.79)** | **1.67** | **(1.01, 2.75)** |
| Relative humidity | 0.762 | 0.952 | 2-6 days | **1.11** | **(1.01, 1.22)** | 1.08 | (0.95, 1.23) |
|  |  |  | 6-14 days | **1.43** | **(1.22, 1.68)** | **1.38** | **(1.11, 1.72)** |
|  |  |  | 14-21 days | 0.90 | (0.78, 1.05) | 1.06 | (0.86, 1.31) |
| Precipitation | 0 mm | 10 mm | 2-6 days | **1.11** | **(1.01, 1.21)** | 1.05 | (0.93, 1.19) |
|  |  |  | 6-14 days | **1.47** | **(1.25, 1.73)** | 1.21 | (0.98, 1.49) |
|  |  |  | 14-21 days | 1.04 | (0.88, 1.21) | 1.01 | (0.82, 1.24) |
| Vapour pressure* | 9.2 hPa | 18.1 hPa | 2-6 days | 1.02 | (0.87, 1.20) | 1.00 | (0.84, 1.19) |
|  |  |  | 6-14 days | **1.59** | **(1.22, 2.08)** | **1.52** | **(1.15, 2.01)** |
|  |  |  | 14-21 days | **1.30** | **(1.01, 1.68)** | **1.31** | **(1.00, 1.71)** |
| Wind speed | 0 m/s | 20 m/s | 2-6 days | 0.97 | (0.72, 1.31) | - |  |
|  |  |  | 6-14 days | 0.99 | (0.59, 1.67) | - |  |
|  |  |  | 14-21 days | 0.87 | (0.53, 1.43) | - |  |
| Maximal gust | 0 m/s | 20 m/s | 2-6 days | 0.98 | (0.90, 1.06) | 0.95 | (0.85, 1.05) |
|  |  |  | 6-14 days | 1.03 | (0.90, 1.19) | 0.97 | (0.82, 1.16) |
|  |  |  | 14-21 days | 0.99 | (0.86, 1.13) | 1.04 | (0.88, 1.23) |
| Atmospheric pressure | 964.6 hPa | 986.8 hPa | 2-6 days | 0.96 | (0.78, 1.18) | 0.89 | (0.70, 1.15) |
|  |  |  | 6-14 days | **0.59** | **(0.42, 0.83)** | 0.70 | (0.47, 1.05) |
|  |  |  | 14-21 days | 1.00 | (0.70, 1.42) | 0.94 | (0.62, 1.42) |

\*Model 2 instead of model 1

Figure 5 shows the lag structure of 21 days for each weather variable in both the single-exposure and multi-exposure model (left-hand panel), and the cumulative odds ratio of the multi-exposure model for the different lag periods (right-hand panel). For example, Figure 5D shows the DLNM outputs for daily mean vapour pressure. The left-hand panel depicts the OR for an increase from 9.2 to 18.1 hPa across lags 0 to 21. Both the single-exposure and multi-exposure model show that vapour pressure has increased ORs 7 to 17 days before disease onset. The right-hand panel depicts the cumulative OR for each lag period ‘2-6 lag days before disease onset’, ‘6-14 lag days’ and ‘14-21 lag days’. The strongest associations can be seen for 6-14 lag days before the exposure, where the OR is around 1.3 (CI: 1.1-1.4) at 15 hPa daily mean vapour pressure.

**Figure 5.**
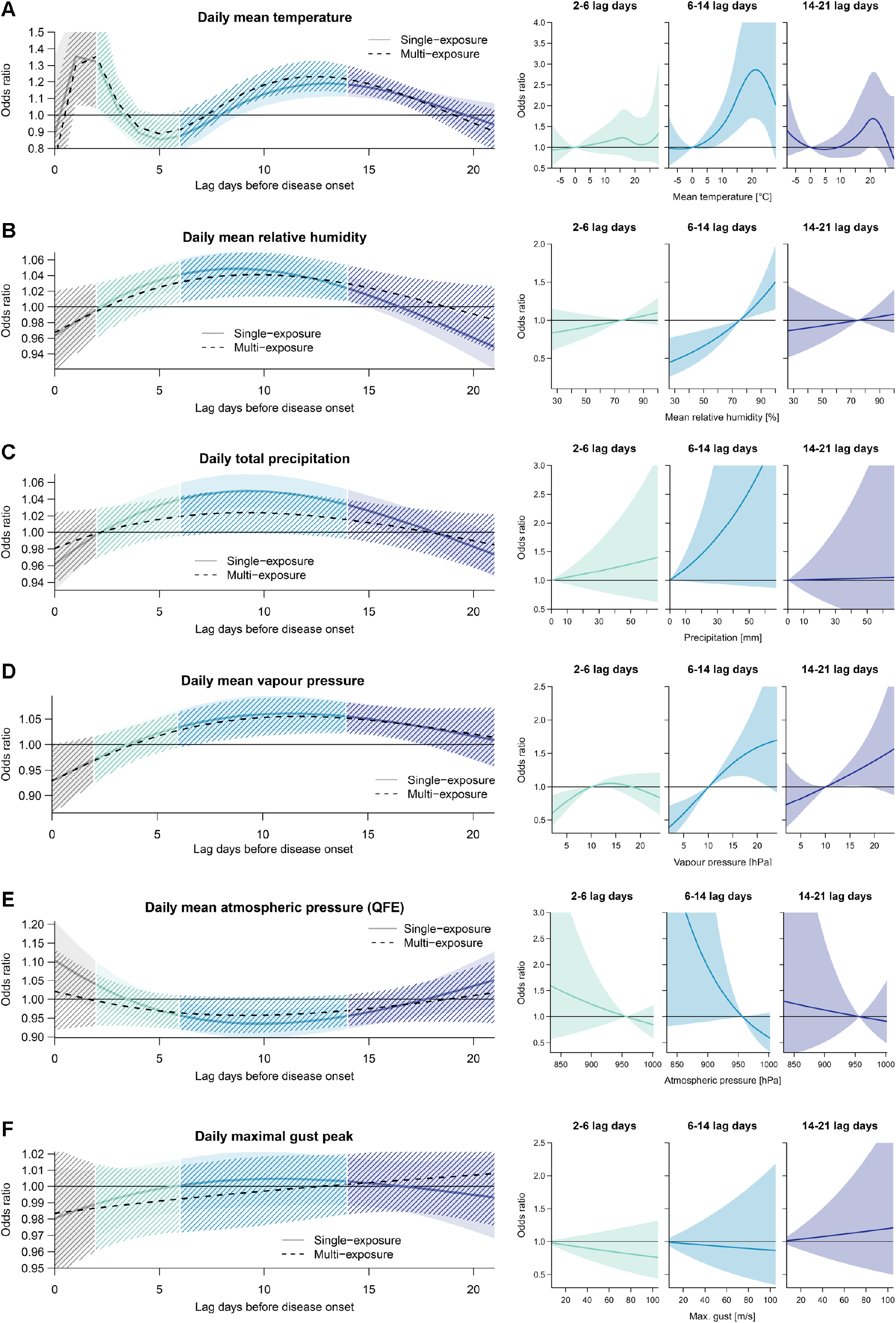
Short-term associations of seven weather variables with LD onset (2017-2021). DLNM model output for (A) daily mean temperature, (B) daily mean relative humidity, (C) daily total precipitation, (D) daily mean vapour pressure, (E) daily maximal gust peak and (F) daily mean atmospheric pressure (QFE). The right-hand figure always depicts the lag structure across 21 days before the Legionnaires’ disease onset. The left-hand figure depicts the overall odds ratio (OR) for three exposure windows: early incubation (lag 2-6), late incubation (lag 6-14) and before incubation (lag 14-21). The multi-exposure models included daily mean relative humidity, daily total precipitation, daily maximal gust peak and daily mean atmospheric pressure (QFE) and either daily mean temperature or daily mean vapour pressure, as well as a term adjusting for regional school holidays.

The mean temperature showed a significantly increased OR at 20 °C compared to the baseline at 0 °C (OR 1.58, CI: 1.05, 2.37) shortly before the disease onset (lag 0-2 prior to first symptoms). Followed by stronger and longer lasting associations at the higher end of the incubation period (lag 6-14) and before the incubation period (lag 14-21) (Figure 5A). Vapour pressure showed the largest OR in the single-exposure models in the 6 to 21 days before onset of the disease. Relative humidity and precipitation were significantly associated with LD incidence during the whole incubation time. Atmospheric pressure was only found statistically significant late in the incubation time (6-14 lag days) and the only weather variable that was negatively associated with LD relative risk.

The estimated direction of association remained consistent in the multi-exposure model. Mean temperature showed a large increase in effect size (OR 2.83, CI: 1.70-4.70), while precipitation and atmospheric pressure were no longer significantly associated.

#### 3.3.3 Sensitivity analysis including daily mean NO_2_ (2019)

Only for the year 2019, data for both, LD case residential addresses and NO_2_ concentrations, were available. Therefore, for a subset of 426 LD cases, the associations with air pollution could be analysed using the same methodology as for the weather variables.

With data restricted to 2019, the analysis (including weather) does not have enough data points to lead to a conclusive result and should be interpreted with caution (Table S5). NO_2_ was most strongly correlated with temperature (r = -0.4) and vapour pressure (r = -0.36). In the single-exposure models, only relative humidity and precipitation remained significant with relative humidity having an OR of 1.23 (CI: 1.00, 1.52) 2-6 lag days before disease onset and an OR of 1.75 (CI: 1.24, 2.47) 6-14 days before disease onset. Ten millimetres of precipitation corresponded to an OR of 1.69 (CI: 1.12, 2.54) 6-14 days before disease onset. The direction of association, however, remained consistent with the analysis of the full dataset. The 95% percentile of NO_2_ (37.3 µg/m^3^) was negatively associated, though statistically not significant, during the incubation time, but positively associated before the incubation, compared to the median (16.5 µg/m^3^).

In the multi-exposure model, the association of relative humidity and precipitation with LD diminished, yet atmospheric pressure showed an OR of 0.29 (CI: 0.10, 0.89) for the 95^th^ percentile compared to the median. NO_2_ was consistently but non-significantly associated with LD occurrence (**Error! Reference source not found.Error! Reference source not found**.).

#### 3.3.4 Sensitivity analysis excluding all cases from Ticino (2017-2021)

The sensitivity analysis excluding Ticino (1,760 LD cases) showed largely consistent estimates with the full data analyses. The largest change was the increasing association of precipitation with LD incidence in the multi-exposure model at lag 6-14 with a 58% (CI: 19-110%) increase in the odds in LD notification per 10 mm increase in daily precipitation (Table S6).

## 4 Discussion

In this study, we provide a comprehensive overview of the spatial distribution and the environmental determinants of LD cases in Switzerland for the years 2017-2021. Overall, our models supports the notion that a specific sequence of weather events - warm weather followed by high humidity, leads to the highest risk for contracting LD^15,18,19^. Understanding the impact of weather on infectious diseases, such as LD supports the interpretation of regional distribution or seasonality of disease. It also opens opportunities for climate- and weather driven early warning systems^47^ and could guide diagnostic testing and treatment preferences for pneumonia patients.

### 4.1 The impact of weather on Legionnaires’ disease incidence

Overall, temperature, relative humidity and vapour pressure were associated with the highest risk increase on case occurrence. These observations are in line with previous studies identifying wet and warm weather to be associated with LD case occurrence^9-11,15^. In particular, our findings confirm the observations of Conza and colleagues from 2013, who analysed the weather conditions in relation to LD incidence of two Swiss regions aggregated by month^20^. Our study contributes to translate these local conclusions to the national level and by using DLNM models allows a more detailed understanding of the time-lagged response of the different weather variables.

Weather can affect case numbers in various ways, such as (i) increasing the susceptibility of the population towards an infection with *Legionella* spp. on the long term; (ii) increasing the current transmission rates through increased bacteria concentrations in the environment or increased exposure (e.g. more air droplets or change in behaviour) or (iii) worsening of symptoms, which results in increased health-seeking, detection and reporting. The ‘worsening of symptoms scenario’ is expected to present the most immediate impact on LD notifications (increased OR on the case day or shortly before). An increased transmission rate could result in an increased OR during or before the incubation period, when the bacteria start to proliferate in the environment. An increase in susceptibility would likely show larger ORs during the incubation period or on the longer term. Therefore, the specific (lag) days for which an association with LD incidence is observed can provide meaningful insights into the mechanisms by which weather affects the LD case occurrence. To date, these dynamics are poorly understood.

Temperature was found to have the strongest association with disease onset early in the incubation period and just before. Consistent with our study, Ricketts et al. observed an increased risk of LD associated with increasing temperatures with long lag periods, up to three months preceding LD infections, which is much longer than our investigated time frame^15^. This would align with the hypothesis that sustained high temperature can warm up small water sources up to *Legionella*’s ideal growth temperatures of 25-45 °C. We observed the highest OR at 21-22 °C during and before the early incubation period. Other studies also found a maximal association at this temperature range^12,19^ that is below *Legionella*’s ideal growth temperature. However, the mean daily temperature used in our study is measured in the ambient air 2 m above ground and might not represent the actual temperatures in e.g. plumbing and piping. Additionally, higher ambient temperatures, i.e. over 24 °C seem to reduce airborne bacterial survival^48^.

We also observed an increased association of elevated temperature with LD case occurrence just before the case date, which is likely too close to the disease onset to fall into the incubation time. The same association has also been reported by others^49^. These short-term relationships have rarely been investigated, but could be explained by a worsening of symptoms in previously infected people due to hot weather - the highest OR was seen at a mean daily temperature of 28 °C likely prompting people to seek care and leading to LD case detection. Mean daily vapour pressure showed similar, yet slightly weaker associations than temperature. Vapour pressure is highly correlated with temperature; it is, therefore, difficult to disentangle the individual contributions of these two weather variables.

Relative humidity showed the strongest association with LD incidence throughout the incubation time, consistent with the existing literature^11,20^. This shorter lag period compared to temperature suggests that humidity may indeed increase LD transmission rates. Daily total precipitation was also found to be linearly associated with LD occurrence in the single-exposure model. Also consistent with our findings, heavy rainfalls were previously found to be associated with LD occurrence^50^. However, the association between precipitation and LD incidence reduced in the multi-exposure model. It is likely that the estimated risk of precipitation may be confounded by daily mean relative humidity. In turn, the association of relative humidity with LD remained significant in both, the single-exposure and multi-exposure models.

Lastly, we found that increasing atmospheric pressure was associated with a decrease in the odds of LD infection in the single-exposure model but not in the multi-exposure model. This association is likely confounded by humidity, which lowers atmospheric pressure^13^. Furthermore, lower atmospheric pressure is also associated with more storms and precipitation. These findings highlight the strong interconnection of the different weather variables, which, in turn, complicate effect attribution to specific weather variables. It is interesting, however, that another Swiss study found only vapour pressure to be associated with LD cases but not relative humidity^20^. We found a significant association for all three in the multi-exposure models: temperature, relative humidity and vapour pressure. While we did not investigate the cumulative effect of different weather variables, weather types, i.e. a combination of weather conditions might be the most suitable predictor for LD incidence in practice.

### 4.2 The role of air pollution

Given the sparse literature on the association of air pollution with LD, we aimed to include air pollution in the case-crossover study as we found strong association with LD case numbers per district for both NO_2_ and PM_2.5_ in the ecological model.

We did not find any significant association between daily air pollution and LD incidence, using a highly resolved spatiotemporal NO_2_ model to estimate exposure. This finding is in part due to a lack of power, as only data from 2019 could be investigated. Even other investigated weather conditions, such as temperature, that showed statistically significant results using data of five years remained inconclusive when only using data of a single study year. Additionally we could only use NO_2_, as a proxy for overall air pollution. Even though PM_2.5_ and NO_2_ showed a similar strength of association in the ecological model, other pollutants (PM or ozone) may play a more important role in LD incidence than NO_2_.

However, we found that the associations between NO_2_ and LD cases in the multi-exposure model were stronger at the beginning of the 21-day period under study than immediately before the onset of the disease. Coupled with the large and significant association observed in the aggregated analysis of the ecological model, this finding could suggest that NO_2_ has less of a short-term or transient effect on LD incidence, but may modify the risk to contract LD in the long-term. Such long-term effects cannot be captured using our case-crossover design. Overall, long-term exposure to NO_2_ could affect the susceptibility to pneumonia in general, for example through the epithelial cell damage or other biological mechanism^51^. Additionally, long-term exposure could increase disease severity, and lead to hospitalisation and thereby case detection and notification. Yet, the strong observed association of air pollution with LD incidence in the ecological model could also stem from an unknown confounder, which occurs primarily in Ticino, as this canton has both the highest air pollution and the highest LD notification rates in Switzerland^35^. Such confounding was, however, avoided in our case-crossover analyses.

Based on the large body of evidence of the impacts of weather and air pollution on human health^52^, we recommend that future studies continue including air pollution in the assessment of weather events on LD incidence with a larger time series, and several pollutants including NO_2_, PM and ozone.

### 4.3 Topography and the regional distribution of Legionnaires’ disease in Switzerland and abroad

The Ticino region in the South of Switzerland shows the highest notification rate of all regions and has been marked a hot spot in the spatial hot spot analysis. However, while the notification rate of LD has been consistently elevated in Ticino and the discrepancy grew stronger in 2015, in recent years the notification rate in Ticino declined, contrary to the rates in the other regions across Switzerland^53^.

The weather in Switzerland is characterised by the Alps dividing the country, leading to strong weather differences even within our small country. In the ecological model, we saw a negative association of relative humidity and LD incidence, which contrasted with the existing literature^11,13,15,16^. In the case-crossover study however, our results concurred well with the available literature. This discrepancy between both study designs could be explained by regional confounding in the ecological model using aggregated data at the regional level, which is absent in the case-crossover design. Since the alpine regions have an overall higher humidity than the rest of Switzerland, but also the lowest notification rates, this particular topography of Switzerland is likely to have driven this unexpected negative association in the ecological model.

The higher LD incidence in Ticino could also be explained by the higher humidity in this region, together with the more frequent occurrence of heavy rainfall events and warmer temperatures. Ticino also lies in the area of influence of the Italian Po-valley with one of the highest air pollution measurements in Europe, particularly for PM and ozone^54^. Whether this special geographical and environmental situation explains the LD hotspot in Ticino will need further inquiry alongside the assessment on the general effects of air pollution on LD using fine-scale air pollution data over several years.

Weather phenomena and air pollution as potential drivers of LD incidence should lead to cross-border effects on the notification numbers of LD. While Switzerland stands out with higher notification rates than the adjacent neighbouring countries, the clustering of higher rates lies towards the area south of the Alps. In addition, Switzerland’s case numbers are not reported through the European Surveillance System managed by the ECDC and are, therefore, missing in the annual epidemiological reports. Looking at the newest report of 2020, only Slovenia, being also part of the alpine belt, had a higher notification rate than Switzerland^55^. Part of this variation is probably depend on health systems factors, such as in Southern Italy where underdiagnosing and underreporting have been previously reported^56,57^.

Since extreme weather conditions such as increasing warm weather and heavy rain events are expected to become more frequent^58^, LD infections are likely to increase in future years. It is, therefore, important that the drivers of LD and their interactions are being understood, especially focussing on the interplay of various correlated weather conditions and air pollution. Future environmental and public health policies focussing on the mitigation of air pollution, together with effective climate actions will be essential to reduce the burden of non-communicable diseases, but also of infectious diseases such as LD.

### 4.4 Strengths and limitations

The combination of several analytical approaches compensated for limitations of a single study design. The ecological design is useful to understand geographical distributions and LD clusters over time, the association between long-term environmental exposures and case numbers might be confounded by further regional, geographic or topographic characteristics, such as health systems performance or altitude. The case-crossover study removed between-individual exposure variability and potential confounding through time-invariant characteristics. DLNMs are particularly useful to study the association of weather with LD incidence due to the models’ ability to investigate sequences and different time delays (lags)^10^. Yet, while DLNMs were found to be well suited for these types of analyses, they might be subject to overfitting. To validate our model fit, we built ordinary simple models aggregating over the significant lag days and using a conditional logistic regression.

The case-crossover study design required the assumption that the place of residency is the source of infection for all community-acquired LD cases. However, the spatial scale of meteorological variables limits the exposure misclassification. Further, while the case-crossover design inherently takes into account time-invariant confounders, time-variant confounders need to be specifically adjusted for. While we did include a term in the model to approximate changes in LD incidence due to travels, we cannot exclude that other varying environmental factors may influence our results.

The incubation time of up to 14 days for LD is rather long. Adding some additional time to account for changes in the environment before the incubation the time under investigation was expanded to 21 days. With such a long timeframe, meaningful associations could be diluted. Therefore, we decided to group the 21 days into three periods: ‘early incubation/ shortly before disease onset’, ‘prolonged incubation period’ and ‘before incubation’. While the grouping does influence the presented numbers, the most influential lag days can be visually identified from the DLNM results.

### 4.5 Conclusion

Our study based on individual weather estimates with high resolution confirms that weather conditions such as warm temperature and increasing humidity are likely to increase the risk of LD case occurrence. At the same time, Switzerland’s summers are setting repeatedly new temperature records and the number of rainstorms seems to be increasing. Against the backdrop of climate change, there is a high risk that the burden of LD will aggravate in the future years. Future research should aim to disentangle the main drivers of LD occurrence. In particular, understanding the interplay of temperature and humidity with air pollution and other regional characteristics could explain hotspots of infection and provide guidance on measures to prevent the conducive effect of warm and humid weather on LD incidence. Environmental policies to combat air pollution and climate change must be afforded due consideration in order to limit the progression of LD infections in Europe in the coming years.

## Supporting information

Supplementary file including additional figures and tables

## Data Availability

All data produced in the present study are available upon reasonable request to the authors and with agreement of the respective data holders.

## Acknowledgments

We thank the Federal Offices (Federal Office of Public Health, Federal Office for the Environment, Federal Office of Energy, Federal Office for Agriculture, Federal Statistical Office, Federal Office of Topography and the Federal Office of Meteorology) and all third parties for providing data. The data that support the findings of this study are available on request from the corresponding author with permission of the respective data holders.

We further thank Benjamin Flückiger (Swiss TPH) for support in the air pollution data acquisition and general R support and Melina Bigler (Swiss TPH) for reviewing the manuscript. This study was funded by the Federal Office of Public Health (contract number 142003961).

