## Supplementary file including additional figures and tables for "Impacts of weather and air pollution on Legionnaires’ disease in Switzerland: a national case-crossover study"

### 1 Supplementary material

**Table S1** Overview of different exposure data sources and resolution included in the ecological model.

| Dataset | Source | Year | Resolution |
|---|---|---|---|
| Compost facilities | CVIS, FOEN | 2020 (CVIS), 2013 (FOEN) | Number per district |
| Land use | FSO | 2004 | Percentage covered by agriculture, industry, settlement, unproductive area |
| Weather | MeteoSwiss | 2017-2020 | 3-month mean of June, July, August by district |
| WWTP | FOEN | 2020 | Number per district |
| SwissTLM | swisstopo | 2016 | Number per district |
| | | | Total shoreline length (minus lake islands) |
| | | | Total river length (over ground) |
| Population density | | | |
| …per district | FSO | 2018 | 1,000 people per km$^2$ |
| …per settled area t | FSO | 2018 | 1,000 people per km$^2$ settled area within distrcit |
| Age of population | FSO | 2018 | Mean population age per district |
| Swiss-SEP | SNC | 2017 | Mean Swiss-SEP per district[1] |
| Air pollution | Meteotest | 2017 | Population-weighted mean $PM_{2.5}$ and $NO_2$ concentrations per district based on 200m$^2$ grid |
| Urbanisation | FSO | 2017 | Categories: densely populated areas, intermediate density areas and sparsely populated areas[2] |

CVIS: Composting Inspectorate
FOEN: Federal Office for the Environment
FSO: Swiss Federal Statistical Office
MeteoSwiss: Federal Office of Meteorology and Climatology
SEP: Socioeconomic position
SNC: Swiss National Cohort
SwissTLM: Swiss topographic landscape model
WWTP: Wastewater treatment plants

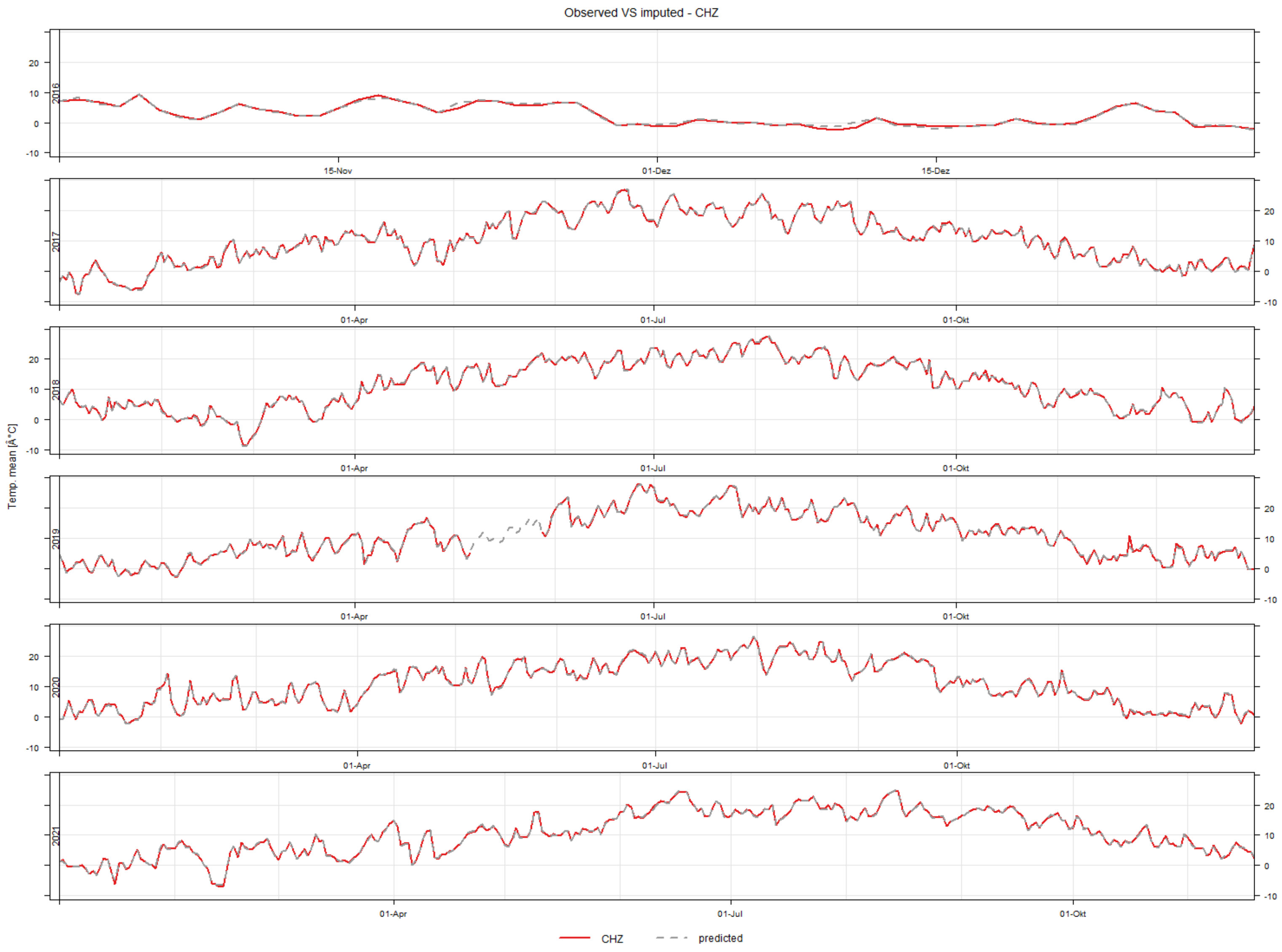


**Figure S1** Illustrative example of the measured (continuous red line) versus imputed (dotted grey line) values of mean temperature over time (Nov 2016-Nov 2021) at one monitoring station (CHZ).

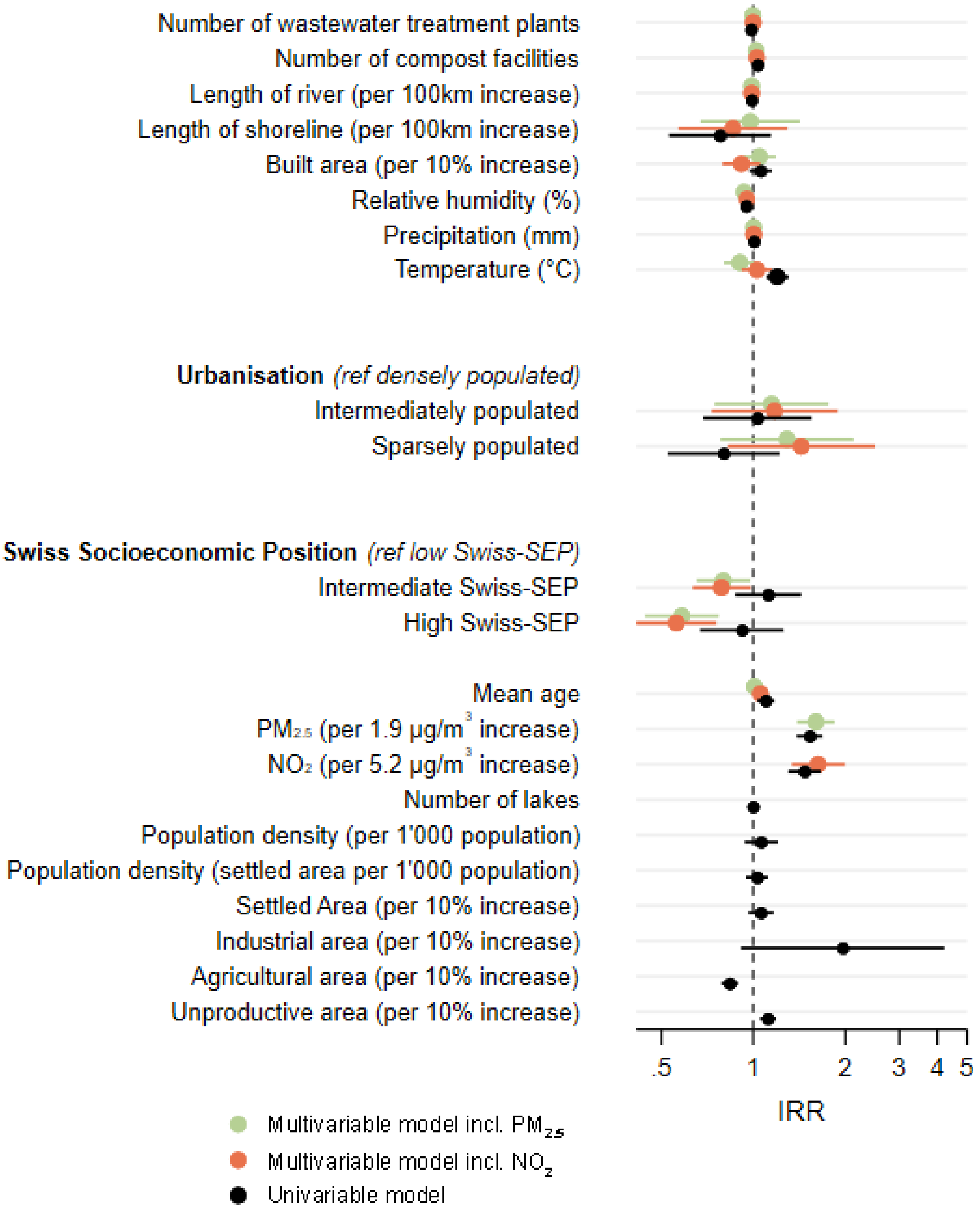


**Figure S2** Incidence rate ratios (IRR) of the univariable and multivariable negative binomial regression analyses of exposure sources and determinants on Legionnaire's disease occurrence per district, 2017-2020 (N=1,603).

**Table S2** Summary statistics of the data availability and model performance for missing data imputation for each parameter for the case-crossover analysis, 2017-2021.

| **Parameter** | **Mean $R^2$** | **Min. $R^2$** | **Max. $R^2$** | **Mean $aR^2$** | **Min. $aR^2$** | **Max. $aR^2$** | **Predictor stations (N)** | **Stations to predict (N)** | **Data availability threshold (%)** | **Total stations (N)** |
|---|---|---|---|---|---|---|---|---|---|---|
| Temperature (max) | 0.991 | 0.968 | 0.998 | 0.990 | 0.966 | 0.997 | 82 | 24 | 80 | 106 |
| Temperature (mean) | 0.996 | 0.990 | 0.999 | 0.996 | 0.990 | 0.999 | 81 | 25 | 80 | 106 |
| Temperature (min) | 0.987 | 0.978 | 0.996 | 0.987 | 0.977 | 0.996 | 82 | 24 | 80 | 106 |
| Rel. humidity (max) | 0.728 | 0.336 | 0.884 | 0.719 | 0.314 | 0.880 | 55 | 51 | 80 | 106 |
| Rel. humidity (mean) | 0.938 | 0.883 | 0.971 | 0.935 | 0.877 | 0.970 | 79 | 27 | 80 | 106 |
| Rel. humidity (min) | 0.907 | 0.817 | 0.962 | 0.902 | 0.809 | 0.960 | 79 | 27 | 80 | 106 |
| Precipitation | 0.837 | 0.681 | 0.949 | 0.833 | 0.673 | 0.948 | 41 | 130 | 75 | 183 |
| Vapour pressure | 0.993 | 0.979 | 0.997 | 0.993 | 0.978 | 0.997 | 78 | 28 | 80 | 106 |
| Wind speed | 0.886 | 0.755 | 0.964 | 0.881 | 0.742 | 0.962 | 82 | 25 | 80 | 108 |
| Max. gust | 0.780 | 0.405 | 0.909 | 0.774 | 0.389 | 0.907 | 43 | 64 | 80 | 108 |
| Atmospheric pressure | 0.999 | 0.990 | 1.000 | 0.999 | 0.989 | 1.000 | 90 | 10 | 80 | 100 |
| **Summary statistics** | | | | | | | | | | |
| Mean | 0.913 | 0.798 | 0.966 | 0.910 | 0.791 | 0.965 | | | | |
| Min | 0.728 | 0.336 | 0.884 | 0.719 | 0.314 | 0.880 | | | | |
| Max | 0.999 | 0.990 | 1.000 | 0.999 | 0.990 | 1.000 | | | | |

**Table S3** Descriptive statistics for weather conditions at station level in Switzerland from 1 November 2016 to 19 November 2021. The table shows the completed dataset using measured and imputed values. As summer, we defined June to August, and for winter December to February.

| | **Central Switzerland** | | **Eastern Switzerland** | | **Espace Mittelland** | | **Lake Geneva Region** | | **Northwestern Switzerland** | | **Zurich** | | **Ticino** | |
|---|---|---|---|---|---|---|---|---|---|---|---|---|---|---|
| | Summer | Winter | Summer | Winter | Summer | Winter | Summer | Winter | Summer | Winter | Summer | Winter | Summer | Winter |
| **Daily mean temperature [ °C] (N=105)** | | | | | | | | | | | | | | |
| Mean (SD) | 17.7 (4.1) | 1.2 (4.3) | 18.1 (3.7) | 0.9 (4.4) | 17.3 (4.0) | 1.0 (4.4) | 17.8 (3.9) | 1.0 (4.5) | 19.6 (3.2) | 2.6 (3.9) | 18.5 (3.7) | 1.64 (4.2) | 18.7 (4.8) | 1.8 (4.4) |
| Median (Min, Max) | 18.0 (1.0, 28.1) | 1.2 (-21.7, 17.2) | 18.3 (2.8, 29.5) | 0.9 (-21.0, 18.2) | 17.0 (1.7, 28.6) | 1.1 (-20.6, 15.9) | 17.9 (4.7, 28.9) | 1.2 (-18.7, 16.6) | 19.6 (10.0, 29.1) | 2.2 (-10.5, 13.8) | 18.6 (7.0, 27.8) | 1.5 (-15.3, 13.8) | 19.6 (2.9, 29.6) | 2.2 (-18.1, 15.1) |
| **Daily mean relative humidity [%] (N=105)** | | | | | | | | | | | | | | |
| Mean (SD) | 75.9 (11.1) | 83.3 (14.2) | 73.4 (11.5) | 80.3 (15.3) | 74.3 (11.3) | 82.2 (14.6) | 69.3 (11.9) | 77.9 (14.7) | 71.7 (11.5) | 84.3 (10.8) | 71.2 (12.3) | 81.9 (13.2) | 70.2 (13.1) | 67.8 (20.7) |
| Median (Min, Max) | 75.9 (34.5, 100.0) | 86.4 (9.2, 100.0) | 73.8 (27.0, 100.0) | 83.3 (14.1, 100.0) | 74.4 (30.8, 100.0) | 85.7 (6.2, 100.0) | 68.90 (24.1, 100.0) | 80.9 (14.1, 100.0) | 71.8 (36.5, 100.00) | 86.2 (38.3, 100.0) | 71.3 (40.8, 100.0) | 84.5 (14.1, 100.0) | 70.7 (24.9, 100.0) | 70.3 (10.6, 100.0) |
| **Daily total precipitation [mm] (N=162)** | | | | | | | | | | | | | | |
| Mean (SD) | 5.3 (9.9) | 3.3 (7.2) | 4.3 (9.1) | 2.8 (6.7) | 3.9 (8.0) | 2.9 (6.1) | 3.1 (6.8) | 3.0 (7.0) | 3.3 (6.9) | 2.8 (5.7) | 4.0 (8.4) | 2.9 (6.1) | 5.4 (15.7) | 2.2 (7.8) |
| Median (Min, Max) | 0.1 (0, 9.8) | 0 (0, 96.3) | 0 (0, 106.5) | 0 (0, 128.6) | 0 (0, 82.3) | 0 (0, 79.4) | 0 (0, 8.6) | 0 (0, 116.2) | 0 (0, 72.5) | 0 (0, 66.0) | 0 (0, 71.1) | 0 (0, 78.9) | 0 (0, 185.4) | 0 (0, 93.5) |
| **Daily mean vapour pressure [hPa] [N=105]** | | | | | | | | | | | | | | |
| Mean (SD) | 15.1 (2.9) | 5.7 (1.7) | 14.9 (3.0) | 5.4 (1.7) | 14.4 (2.8) | 5.5 (1.8) | 13.8 (2.9) | 5.3 (1.8) | 15.8 (2.6) | 6.4 (1.7) | 14.8 (2.5) | 5.7 (1.6) | 15.2 (4.2) | 4.8 (1.9) |
| Median (Min, Max) | 15.3 (4.2, 23.9) | 5.7 (0.4, 13.2) | 15.1 (5.3, 25.6) | 5.4 (0.7, 13.2) | 14.4 (4.3, 24.4) | 5.50 (0.5, 13.6) | 13.8 (4.0, 26.2) | 5.2 (0.9, 12.0) | 15.8 (8.3, 24.0) | 6.1 (1.2, 13.2) | 14.6 (6.9, 23.4) | 5.6 (1.0, 11.5) | 15.3 (3.6, 27.3) | 4.7 (0.5, 11.3) |
| **Daily mean wind speed (scalar) [m/s] (N=107)** | | | | | | | | | | | | | | |
| Mean (SD) | 6.8 (3.1) | 8.1 (6.8) | 7.4 (3.1) | 7.8 (6.1) | 8.0 (5.1) | 10.7 (10.1) | 8.3 (3.8) | 8.7 (6.8) | 5.6 (2.4) | 8.3 (6.0) | 8.7 (4.3) | 12.0 (8.7) | 8.6 (5.7) | 8.9 (8.3) |
| Median (Min, Max) | 6.1 (1.1, 36.7) | 5.4 (0.4, 51.5) | 6.8 (1.4, 30.2) | 5.8 (0, 49.7) | 6.5 (1.1, 57.2) | 7.2 (0.4, 78.8) | 7.20 (1.8, 46.4) | 6.1 (0, 51.8) | 5.0 (1.1, 18.7) | 6.8 (0.7, 45.7) | 7.6 (1.8, 30.2) | 9.4 (1.1, 58.7) | 6.8 (1.8, 48.6) | 5.8 (0, 60.8) |
| **Daily gust peak (1 second) [m/s] (N=107)** | | | | | | | | | | | | | | |
| Mean (SD) | 33.0 (15.8) | 33.7 (25.2) | 34.7 (13.6) | 32.5 (20.5) | 35.6 (14.9) | 38.0 (25.6) | 36.7 (13.8) | 33.8 (20.5) | 30.8 (12.0) | 33.8 (20.3) | 35.6 (14.9) | 39.8 (23.8) | 36.4 (15.9) | 32.1 (21.6) |
| Median (Min, Max) | 28.8 (6.1, 161.6) | 23.8 (3.6, 195.1) | 32.4 (7.9, 133.2) | 26.3 (0, 139.0) | 32.0 (9.0, 122.8) | 30.2 (5.4, 183.6) | 34.8 (9.0, 129.6) | 28.4 (5.0, 147.6) | 28.1 (8.3, 134.3) | 29.2 (5.4, 147.6) | 32.4 (11.2, 131.8) | 34.6 (6.1, 163.1) | 32.4 (7.6, 122.4) | 25.2 (4.3, 133.6) |
| **Atmospheric pressure (QFA) [hPa] (N=99)** | | | | | | | | | | | | | | |
| Mean (SD) | 931.5 (49.1) | 931.1 (52.5) | 942.8 (41.7) | 943.1 (44.9) | 926.1 (46.3) | 925.4 (49.7) | 918.7 (47.7) | 918.2 (51.1) | 972.5 (11.5) | 974.3 (15.6) | 942.8 (28.2) | 942.8 (31.5) | 921.8 (71.1) | 921.6 (75.7) |
| Median (Min, Max) | 955.8 (776.5, 974.0) | 948.7 (756.6, 988.8) | 960.0 (798.4, 985.6) | 959.0 (780.1, 1002.6) | 948.5 (789.1, 981.2) | 946.1 (768.5, 996.2) | 929.0 (822.1, 980.1) | 931.7 (805.0, 995.1) | 976.3 (929.4, 988.8) | 977.3 (912.2, 1004.0) | 956.3 (873.5, 976.0) | 955.0 (855.1, 990.7) | 956.9 (768.8, 999.0) | 959.9 (750.8, 1017.1) |

**Table S4** Output for simple conditional logistic regression for LD cases 2017-2021. Odds ratios and 95% confidence intervals for single-exposure and multi-exposure models for each weather variable (without $NO_2$). Due to collinearity, two models were constructed, once with temperature and the other with vapour pressure. All estimates stem from the mean temperature model (Model 1), except vapour pressure, which is based on the vapour pressure model (Model 2). The unit increase correspondence to the same increase from "center" to "value", that was used for the DLNM.

| | | **Single-exposure** | | **Multi-exposure** | |
|---|---|---|---|---|---|
| **Parameter** | **Increase** | **OR** | **95% CI** | **OR** | **95% CI** |
| Temperature | 20 °C | 3.07 | (1.92, 4.91) | 2.92 | (1.80, 4.71) |
| Relative humidity | 19% | 1.58 | (1.37, 1.82) | 1.39 | (1.18, 1.64) |
| Precipitation | 10 mm | 1.46 | (1.30, 1.63) | 1.20 | (1.05, 1.37) |
| Vapour pressure* | 8.9 hPa | 2.05 | (1.58, 2.65) | 1.89 | (1.45, 6.56) |
| Wind speed | 20 m/s | 0.94 | (0.61, 1.43) | . | |
| Maximal gust | 20 m/s | 0.94 | (0.84, 1.06) | 0.98 | (0.87, 1.10) |
| Atmospheric pressure | 22.2 hPa | 0.45 | (0.33, 0.62) | 0.64 | (0.46, 0.90) |

*Model 2 instead of model 1

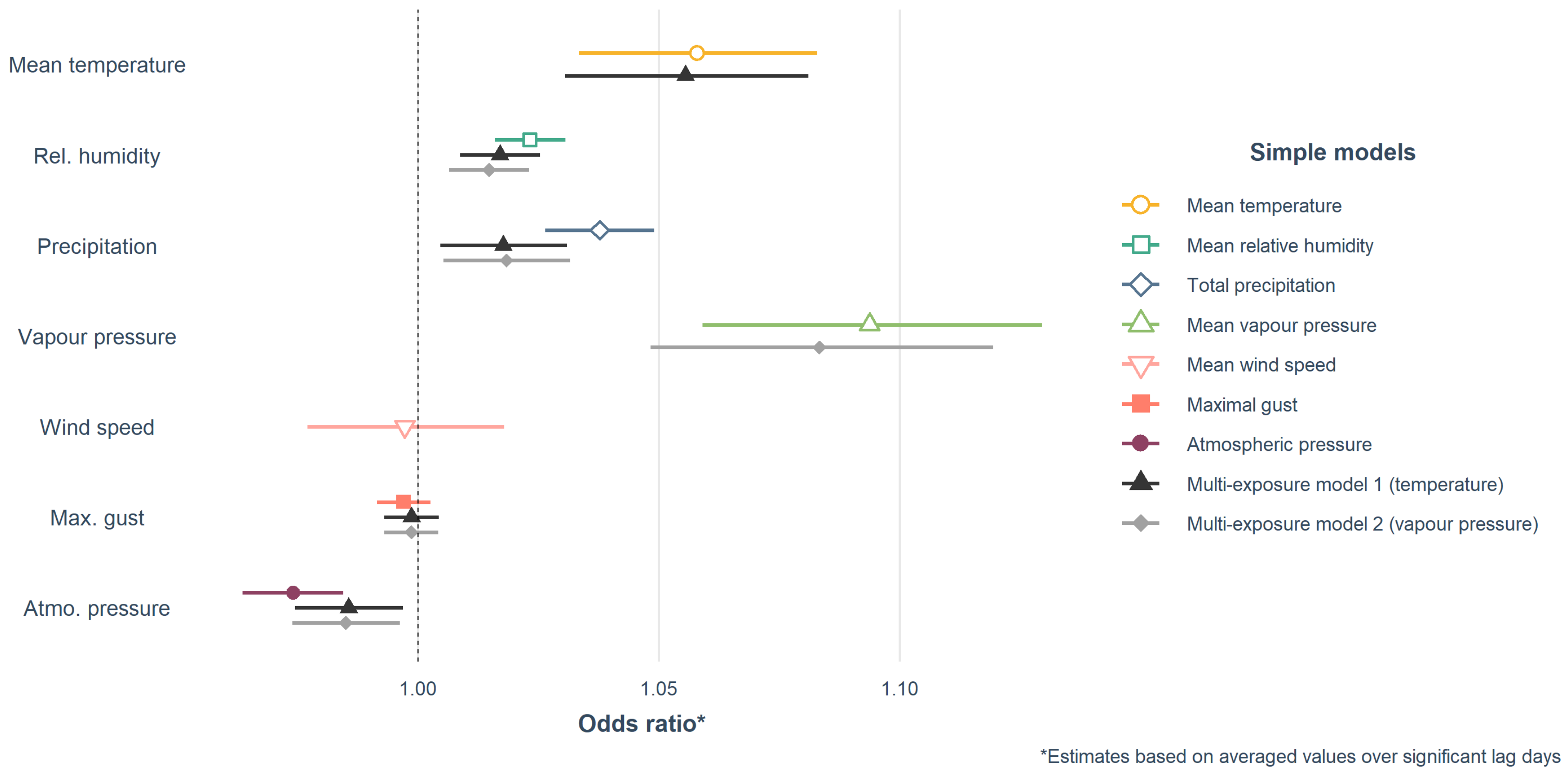


**Figure S3** Forest plot showing the odds ratio and 95% confidence intervals from the single-exposure and both multi-exposure models for a single unit-increase (N=2,047).

**Table S5** Output for DLNM models using conditional logistic regression for LD cases 2019 (N=2,047). Odds ratios and 95% confidence intervals for single-exposure and multi-exposure models for each weather variable for data from 2019 only but in addition of mean daily $NO_2$. Due to collinearity, two models were constructed, once with temperature and the other with vapour pressure. All estimates stem from the mean temperature model (Model 1), except vapour pressure, which is based on the vapour pressure model (Model 2). The centre depicts the reference value selected for the prediction. The value depicts the value for which the overall odds ratio are estimates.

| | | | | **Single-exposure** | | **Multi-exposure** | |
|---|---|---|---|---|---|---|---|
| **Parameter** | **Center** | **Value** | **Lag period** | **OR** | **95% CI** | **OR** | **95% CI** |
| Temperature | 0 °C | 20 °C | 2-6 days | 0.57 | (0.2, 1.63) | 1.22 | (0.34, 4.36) |
| | | | 6-14 days | 1.49 | (0.45, 4.88) | 3.42 | (0.58, 20.29) |
| | | | 14-21 days | 1.03 | (0.31, 3.47) | 2.46 | (0.46, 13.08) |
| Relative humidity | 0.762 | 0.952 | 2-6 days | **1.23** | **(1.00, 1.52)** | 1.05 | (0.77, 1.42) |
| | | | 6-14 days | **1.75** | **(1.24, 2.47)** | 1.17 | (0.68, 2.01) |
| | | | 14-21 days | 1.04 | (0.74, 1.47) | 1.37 | (0.82, 2.30) |
| Precipitation | 0 mm | 10 mm | 2-6 days | 1.04 | (0.82, 1.34) | 1.01 | (0.73, 1.38) |
| | | | 6-14 days | **1.69** | **(1.12, 2.54)** | 1.63 | (0.96, 2.77) |
| | | | 14-21 days | 0.99 | (0.66, 1.49) | 0.81 | (0.45, 1.45) |
| Vapour pressure* | 9.2 hPa | 18.1 hPa | 2-6 days | 0.99 | (0.71, 1.40) | 1.03 | (0.71, 1.50) |
| | | | 6-14 days | 1.13 | (0.61, 2.07) | 1.32 | (0.67, 2.60) |
| | | | 14-21 days | 1.05 | (0.58, 1.91) | 1.35 | (0.68, 2.66) |
| Wind speed | 0 m/s | 20 m/s | 2-6 days | 0.82 | (0.43, 1.59) | . | |
| | | | 6-14 days | 0.58 | (0.19, 1.73) | . | |
| | | | 14-21 days | 0.88 | (0.30, 2.53) | . | |
| Maximal gust | 0 m/s | 20 m/s | 2-6 days | 0.91 | (0.77, 1.07) | 0.92 | (0.72, 1.17) |
| | | | 6-14 days | 0.91 | (0.68, 1.21) | 0.93 | (0.63, 1.38) |
| | | | 14-21 days | 0.97 | (0.73, 1.29) | 1.16 | (0.78, 1.73) |
| Atmospheric pressure | 964.6 hPa | 986.8 hPa | 2-6 days | 1.03 | (0.70, 1.53) | 0.63 | (0.33, 1.18) |
| | | | 6-14 days | 0.59 | (0.33, 1.09) | **0.29** | **(0.10, 0.89)** |
| | | | 14-21 days | 0.87 | (0.44, 1.7) | 0.40 | (0.14, 1.11) |
| $NO_2$ | 16.5 µg/m3 | 37.3 µg/m3 | 2-6 days | 0.72 | (0.44, 1.17) | 1.00 | (0.38, 2.65) |
| | | | 6-14 days | 0.76 | (0.44, 1.34) | 1.58 | (0.52, 4.82) |
| | | | 14-21 days | 1.14 | (0.56, 2.32) | 2.50 | (0.74, 8.43) |

*Model 2 instead of model 1

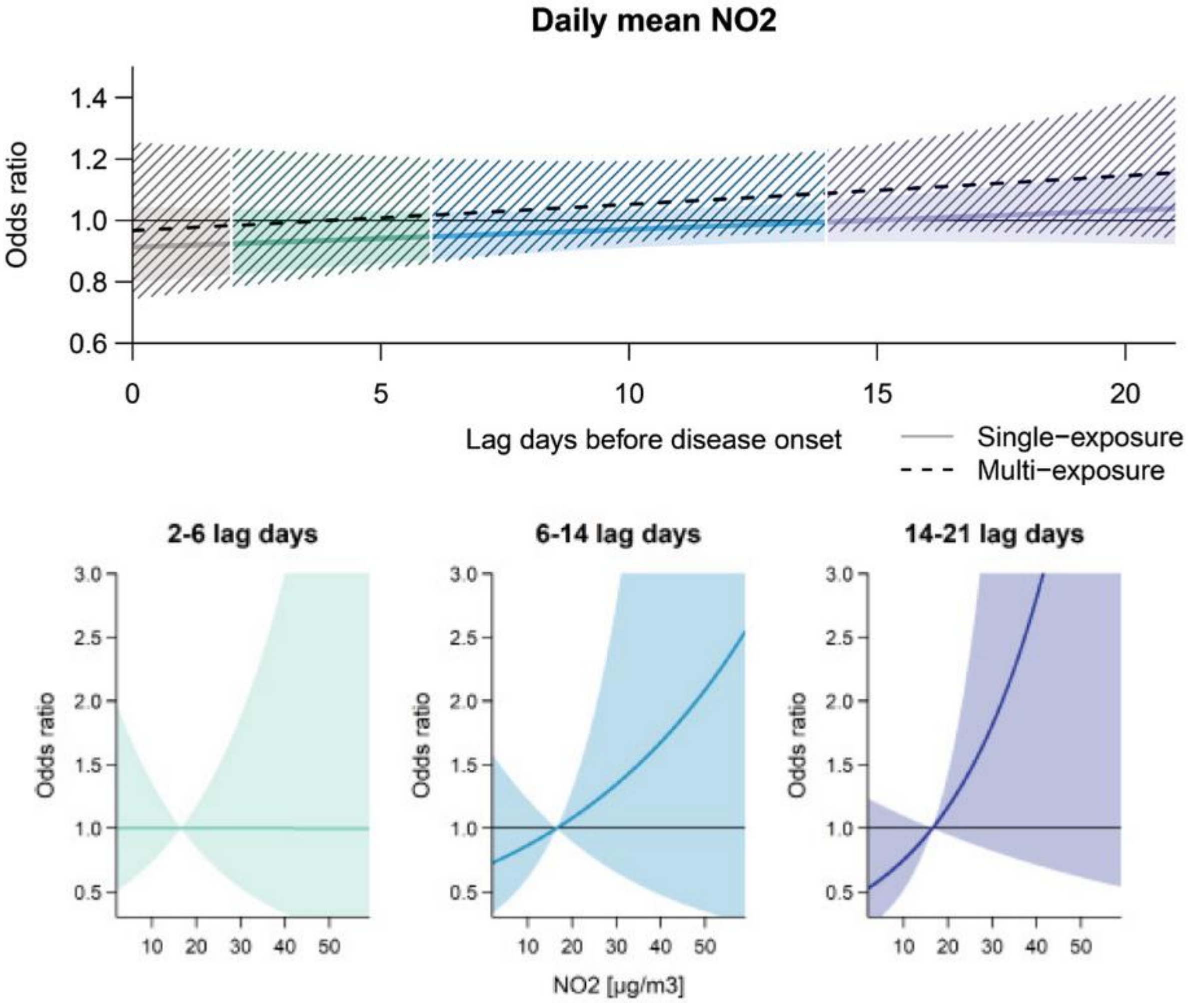


**Figure 4 DLNM model output for daily mean $NO_2$ level for the year 2019** (N=426)**.** The upper figure depicts the lag structure across 21 days before the Legionnaires' disease onset. The lower figure depicts the overall odds ratio (OR) for three exposure windows: early incubation (lag 2-6), late incubation (lag 6-14) and before incubation (lag 14-21). The multi-exposure models included daily mean relative humidity, daily total precipitation, daily maximal gust peak and daily mean atmospheric pressure (QFE) and either daily mean temperature or daily mean vapour pressure, as well as a term adjusting for regional school holidays.

**Table S6 Output for DLNM models using conditional logistic regression for LD cases excluding Ticino 2017-2021** (N=1,760)**.** Odds ratios and 95% confidence intervals for single-exposure and multi-exposure models for each weather variable for all cases excluding Ticino (and without $NO_2$). Due to collinearity, two models were constructed, once with temperature and the other with vapour pressure. All estimates stem from the mean temperature model (Model 1), except vapour pressure, which is based on the vapour pressure model (Model 2). The centre depicts the reference value selected for the prediction. The value depicts the value for which the overall odds ratio are estimates.

| | | | | **Single-exposure** | | **Multi-exposure** | |
|---|---|---|---|---|---|---|---|
| **Parameter** | **Center** | **Value** | **Lag period** | **OR** | **95% CI** | **OR** | **95% CI** |
| Temperature | 0 °C | 20 °C | 2-6 days | 1.22 | (0.76, 1.95) | 1.48 | (0.9, 2.43) |
| | | | 6-14 days | **2.03** | **(1.34, 3.75)** | 3.36 | (1.93, 5.85) |
| | | | 14-21 days | **1.77** | **(1.09, 2.87)** | 1.47 | (0.85, 2.55) |
| Relative humidity | 0.762 | 0.952 | 2-6 days | 1.11 | (0.98, 1.23) | 1.04 | (0.89, 1.21) |
| | | | 6-14 days | **1.52** | **(1.26, 1.83)** | **1.33** | **(1.02, 1.74)** |
| | | | 14-21 days | 0.88 | (0.74, 1.04) | 0.96 | (0.74, 1.23) |
| Precipitation | 0 mm | 10 mm | 2-6 days | **1.20** | **(1.06, 1.36)** | 1.12 | (0.95, 1.32) |
| | | | 6-14 days | **1.92** | **(1.54, 2.38)** | **1.58** | **(1.19, 2.10)** |
| | | | 14-21 days | 1.16 | (0.95, 1.41) | 1.24 | (0.95, 1.62) |
| Vapour pressure* | 9.2 hPa | 18.1 hPa | 2-6 days | 1.12 | (0.97, 1.30) | 1.09 | (0.93, 1.28) |
| | | | 6-14 days | **1.83** | **(1.43, 2.33)** | **1.66** | **(1.28, 2.14)** |
| | | | 14-21 days | **1.27** | **(0.99, 1.63)** | 1.24 | (0.95, 1.61) |
| Wind speed | 0 m/s | 20 m/s | 2-6 days | 0.97 | (0.70, 1.34) | . | |
| | | | 6-14 days | 0.86 | (0.49, 1.51) | . | |
| | | | 14-21 days | 0.93 | (0.55, 1.59) | . | |
| Maximal gust | 0 m/s | 20 m/s | 2-6 days | 0.98 | (0.89, 1.07) | 0.95 | (0.85, 1.07) |
| | | | 6-14 days | 1.00 | (0.86, 1.16) | 0.92 | (0.76, 1.11) |
| | | | 14-21 days | 0.99 | (0.85, 1.15) | 0.98 | (0.82, 1.18) |
| Atmospheric pressure | 964.6 hPa | 986.8 hPa | 2-6 days | 1.04 | (0.86, 1.26) | 0.98 | (0.77, 1.24) |
| | | | 6-14 days | 0.67 | (0.48, 0.91) | 0.76 | (0.52, 1.12) |
| | | | 14-21 days | 0.96 | (0.69, 1.33) | 0.98 | (0.66, 1.46) |

*Model 2 instead of model 1
